# Do standard model assumptions realistically represent HIV dynamics in sex workers? A modelling analysis of South African data

**DOI:** 10.64898/2026.03.10.26348008

**Authors:** Nanina Anderegg, Matthias Egger, Kholi Buthelezi, Yonela Sinqu, Mariette Slabbert, Leigh F. Johnson

**Affiliations:** Department of Infectious Diseases and Hospital Epidemiology, University Hospital Zurich, University of Zurich, Zurich, Switzerland; Institute of Medical Virology, University of Zurich, Zurich, Switzerland; Centre for Integrated Data and Epidemiological Research, School of Public Health, University of Cape Town, Cape Town, South Africa; Population Health Sciences, Bristol Medical School, University of Bristol, Bristol, United Kingdom; SISONKE, National Sex Workers Movement, South Africa; Independent consultant, Pretoria, South Africa

## Abstract

Female sex workers (FSW) in sub-Saharan Africa experience disproportionately high risks of HIV infection. Mathematical models are widely used to assess the contribution of sex workers and other key populations to HIV transmission dynamics and to inform targeted programmes. However, many rely on simplifying assumptions, such as stable sex worker characteristics and constant HIV transmission risk over time. These assumptions may be unrealistic and could bias modelled estimates. We used the South African Thembisa model to assess how alternative assumptions about FSW age, duration of sex work, and client-to-FSW transmission risk affect modelled HIV outcomes. We compared six scenarios that combined constant and increasing FSW age and sex work duration with constant and early-epidemic declining (exponentially or exposure-dependent) transmission risk. Each scenario was calibrated to HIV prevalence data from population-based and sex worker-specific surveys. Scenarios that allowed both FSW characteristics and transmission risk to vary over time showed the best agreement with external data, most closely reproducing HIV incidence, prevalence, and viral suppression estimates from a 2019 national sex worker survey (incidence ∼5 per 100 person-years, prevalence 61-62%, viral suppression ∼60%), and producing incidence rate ratios more consistent with estimates from the broader eastern and southern Africa region. By contrast, the scenario assuming constant FSW characteristics and transmission risk overestimated HIV incidence and underestimated prevalence and viral suppression. At the same time, this time-invariant specification attributed a much larger share of new HIV infections to sex work, with commercial sex work accounting for more than 20% of new infections in 2025, compared with 9-13% under time-varying assumptions. Overall, our findings show that HIV model estimates for sex workers are highly sensitive to modelling assumptions. Incorporating time-varying FSW parameters yields estimates that are more consistent with empirical data and support more reliable programme planning and evaluation.

**Author Summary:** Female sex workers in sub-Saharan Africa face much higher risks of HIV infection than other women. Mathematical models are often used to understand why and to guide prevention programmes. Yet many of these models make simple assumptions about sex workers - for example, that their average age stays the same over time, that they spend a fixed number of years in sex work, or that the chance of HIV passing from a client to a sex worker never changes. In reality, these factors changed over time. In this study, we used South Africa’s national HIV model to test how changing these assumptions affects the results. We compared different versions of the model and checked which ones best matched national sex worker survey data. We found that the model worked better when we allowed sex workers to become older over time, to spend longer in sex work, and the risk of passing on HIV to decline. Our findings show that mathematical models can give very different answers depending on how they represent the lives and experiences of sex workers. More realistic assumptions lead to more accurate estimates and can help ensure that programmes focus support where it is most needed.

## Introduction

Sub-Saharan Africa remains home to most of the world’s people living with HIV (PLHIV), with 65% of global PLHIV residing in the region in 2023 [1]. Because HIV prevalence in the general population is high, the epidemic in this region is often described as “generalized.” As a result, HIV programmes have typically focused on the broader population rather than tailoring interventions to key populations such as sex workers, men who have sex with men, transgender women, or people who inject drugs.

However, even in generalized epidemic settings, key populations face a disproportionately high risk of HIV infection [2, 3]. And even when they account for only a small share of new infections at a given moment, their long-term contribution to HIV incidence, including secondary infections, can be substantial [2–4]. It has also been argued that although general-population approaches have contributed to declining HIV incidence in many African countries, failing to address key populations may undermine long-term epidemic control, with HIV becoming more concentrated in these groups as incidence falls overall [5,6]. Key population prevention and care programmes have been disproportionately affected by the recent cuts in U.S. funding [7].

Female sex workers (FSW) in sub-Saharan Africa are a vulnerable group, with HIV incidence rates roughly eight times average local incidence rates in women of the same age [8]. Approximately 5% of new infections in East and Southern Africa, and 15% of new infections in West and central Africa, occur in sex workers [9]. These estimates depend on mathematical models, which are also used extensively in estimating the impact and cost-effectiveness of sex worker-focused programmes [10–12]. Such models depend on several assumptions about sex workers and are not infallible [13]. Given the critical role of these models in developing HIV programmes, it is important that their assumptions are carefully reviewed.

A common assumption is that the characteristics of sex workers remain stable over time. Yet evidence from South Africa and other settings indicates that the average age of sex workers and duration of sex work have been increasing [14,15]. Another common assumption is that HIV transmission probabilities between sex workers and their clients remain constant over time, after accounting for the HIV-positive partner’s CD4 count, viral load, and the use of prevention methods. However, evidence from Kenya indicates that these probabilities were not stable in the early stages of the epidemic, when per-act transmission rates fell sharply [16]. One possible explanation is a reduction in the prevalence of sexually transmitted infections (STIs), which are known to increase HIV transmission risk [17,18]. Alternatively, more susceptible individuals may have acquired HIV earlier in the epidemic, leaving a population with lower average susceptibility and thus an observed decline in transmission risk. Such heterogeneity in susceptibility could be driven by behavioural differences (e.g., number of clients or condom use) or biological factors (e.g., genetic predispositions or acquired immunity [19]). Regardless of the cause, this variation in per-act transmission risk is not well captured in most HIV transmission models [20,21].

In this study, we use a mathematical of the HIV epidemic in South Africa to simulate HIV dynamics in sex workers under different assumptions about trends in sex worker characteristics and transmission probabilities. South Africa has the largest HIV epidemic in the world [1], with exceptionally high levels of HIV prevalence in sex workers (around 60% in recent surveys [22,23]). Although the country has made substantial progress in reducing HIV incidence overall, incidence remains high among sex workers [24], and there is an urgent need for renewed focus on this group in the wake of recent HIV funding cuts.

## Methods

We used the mathematical model Thembisa to simulate the HIV epidemic in South Africa, with a focus on sex workers and their clients. We varied key assumptions about FSW age distribution, duration in sex work, and per-act client-to-FSW HIV transmission risk, and combined these into a set of alternative scenarios. Each scenario was calibrated to empirical HIV prevalence data and used to estimate HIV incidence, prevalence, viral suppression, incidence rate ratios, and the population attributable fraction of commercial sex work. Model outputs were compared across scenarios and evaluated against external data where available. Further details of the model structure, assumptions, calibration, and analyses are described below.

### Model description

We used version 4.7 of the Thembisa model, a well-established deterministic compartmental model of the HIV epidemic in South Africa that incorporates HIV transmission, disease progression, and the effects of antiretroviral therapy (ART) [25]. Thembisa stratifies the population by demographic and behavioural factors, including high- and low-risk groups defined by the propensity for concurrent partnerships and commercial sex. Here, we focus on the component representing FSW and their clients (Figure 1). FSW are represented as unmarried women in the high-risk group who may enter sex work at age-dependent rates, while men in the high-risk group are assumed to visit sex workers at age- and marital status-dependent rates. The FSW component of Thembisa relies on several parametric assumptions, including assumptions related to condom use in client-FSW contacts, the effect of knowledge of HIV status on entry into sex work, the age distribution of FSW, the duration of sex work, and HIV transmission risk. These assumptions have historically been treated as time-invariant, with fixed parameters over the course of the epidemic. We relaxed this assumption for two FSW characteristics (age and sex work duration) and for HIV transmission risk (Figure 1).

**Figure 1.**
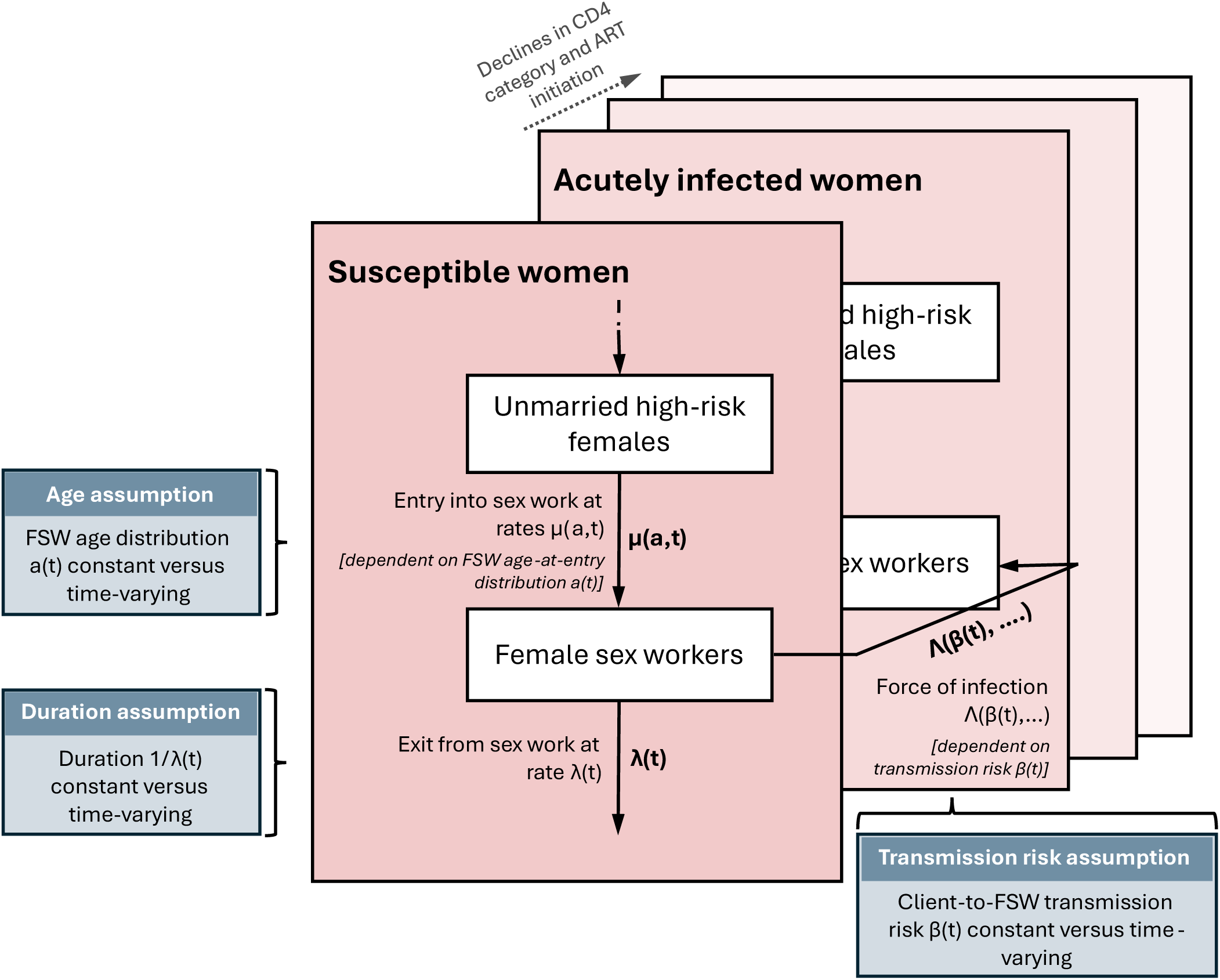
Simplified schematic of the female sex worker (FSW) component of the Thembisa model. Unmarried high-risk women may enter sex work at rates determined by the age-at-entry distribution and exit sex work at a rate determining the average duration of sex work. Susceptible FSW acquire HIV infection through the force of infection, which depends, among other factors, on the client-to-FSW transmission risk. Assumptions regarding the FSW age distribution, duration of sex work, and client-to-FSW transmission risk were varied in this study.

### Assumptions on sex worker characteristics

We considered two specifications for FSW age and duration of sex work: (a) time-invariant distributions consistent with Thembisa v4.7 [25], and (b) time-varying distributions informed by our previous analysis of trends between 1996 and 2019 [14]. In brief, specification (a) assumed a constant mean FSW age of 29 years and mean duration of sex work of 3 years (Supplementary Figure 1). Specification (b) assumed that the mean age increased from 26.4 to 32.2 years and the mean duration of sex work from 2.7 to 7.4 years between 1996 and 2019, remaining constant outside this period (Supplementary Figure 1).

### Assumptions on HIV transmission risk

We considered three specifications for the per-act probability of HIV transmission from an HIV-positive client to a susceptible FSW: (1) a time-invariant risk consistent with earlier versions of Thembisa (≤v4.2), (2) a time-varying specification with an exponential early-epidemic decline as implemented in Thembisa v4.7 [25] and informed by Kenyan data [16], and (3) a more flexible time-varying specification, also informed by Kenyan data [16] and earlier studies [26–28], in which transmission risk declined as a function of the average duration of exposure to HIV-positive clients (Supplementary Figure 1). More details on assumptions about FSW characteristics and HIV transmission risk are provided in Table 1 and <u>Supplementary Text S1</u>.

**Table 1.** Parameter values underlying FSW characteristics and transmission-risk assumptions.

| Parameter | Assumption | Parameter values | Source |
| --- | --- | --- | --- |
| FSW age distribution | Time-invariant (a) | Shifted gamma distribution with offset 10 years and fixed parameters such that:<br>• Mean = 29 years<br>• Variance = 9 | Thembisa version 4.7 default |
|  | Time-varying: increase (b) | Shifted gamma distribution with offset 10 years and time-varying parameters such that:<br>• Mean age increases from 26.4 (1996) to 32.3 (2019)<br>• SD increases from 5.5 to 7.5 | Derived from prior work (Anderegg et al.) |
| Duration of sex work | Time-invariant (a) | Exponential distribution with fixed parameter such that:<br>• Mean duration = 3 years | Thembisa version 4.7 default |
|  | Time-varying (b) | Exponential distribution with time-varying parameters such that:<br>• Mean increases from 2.7 years (1996) to 7.4 years (2019) | Derived from prior work (Anderegg et al.) |
| Client-to-FSW transmission risk | Constant (1) | Constant per-act transmission probability $\tau$<br>• $\tau$ estimated during calibration using Beta prior (mean 0.001, SD = 0.0005). | Based on earlier Thembisa versions ($\leq 4.2$) |
| | Time-varying: exponential early-epidemic decline (2) | Early-epidemic level parameter $R_{\text{FSW-start-epidemic}}$ resulting in higher acquisition risk at the beginning of the epidemic, with **exponential** decay over calendar time toward the baseline transmission probability $\tau$.<br>• $\tau$ (Beta prior) and $R_{\text{FSW-start-epidemic}}$ (Gamma prior) estimated during calibration. | Kenyan data (Kimani et al.), Thembisa version 4.7 |
| | Time-varying: exposure-dependent change (3) | Early-epidemic level parameter $R_{\text{FSW-start-epidemic}}$ resulting in higher acquisition risk at the beginning of the epidemic, with an **exposure**-dependent decay toward the baseline transmission $\tau$<br>• $\tau$ (Beta prior) and $R_{\text{FSW-start-epidemic}}$ (Gamma prior) estimated during calibration.<br>• Decline depends on client HIV prevalence, ART coverage, and duration of sex work. | Kenyan data (Kimani et al.) |

### Scenarios and model fitting

Combining the two specifications for sex worker characteristics with the three specifications for HIV transmission risk produced six scenarios: 1a, 1b, 2a, 2b, 3a and 3b (Table 2). We calibrated each scenario separately using a Bayesian approach, allowing 10 parameters to vary in scenarios 1a and 1b, and 11 parameters in scenarios 2a-3b due to the inclusion of an additional parameter capturing the early-epidemic excess in per-act client-to-FSW transmission risk assumed in the time-varying transmission risk scenarios. The calibrated parameters related primarily to mixing patterns, transmission probabilities, and key aspects of sex worker dynamics (Supplementary Table 1). (Supplementary Table 1).

**Table 2.**
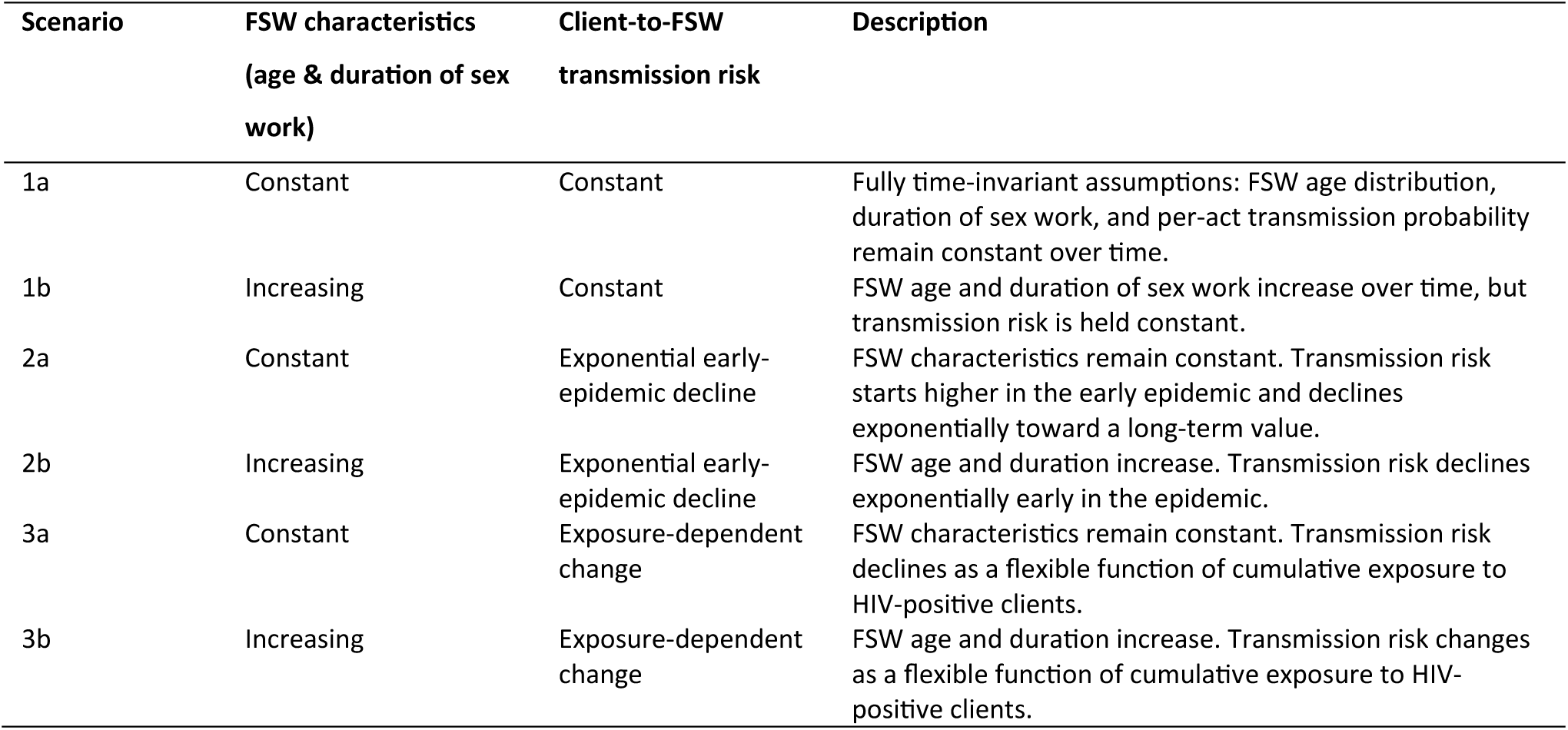
Summary of assumptions about female sex worker (FSW) characteristics and client-to-FSW HIV transmission risk implemented in the six scenarios.

We calibrated each scenario to empirical HIV prevalence data used in the Thembisa v4.7 model calibration framework [25]. These data are based on several long-standing surveillance sources in South Africa, including antenatal clinic (ANC) HIV prevalence surveys conducted annually since 1990 (and less frequently since 2015), age- and sex-specific HIV prevalence estimates from nationally representative household surveys conducted by the Human Sciences Research Council at multiple time points (2005-2022), and HIV prevalence estimates among female sex workers from multiple local surveys conducted since the late 1990s (including the South African Health Monitoring Surveys and earlier city-specific studies) [25].

### Model outcomes

For each of the six scenarios, we estimated time trends from 1985 to 2025 in HIV incidence, HIV prevalence, and viral load suppression among FSW living with HIV. We also estimated age-standardized incidence rate ratios (IRRs), defined as HIV incidence in FSW divided by age-standardized incidence in the general female population, with the latter obtained by weighting annual age-specific incidence rates in women by the annual age distribution of susceptible FSW. In addition, we calculated the population attributable fraction (PAF) of commercial sex work, defined as the proportion of new HIV infections in FSW and in their clients (restricted to infections acquired through contact with FSW) among all new HIV infections each year. Beyond outcomes in FSW, we examined HIV prevalence in the general population and among pregnant women to compare model estimates with calibration data. For all outcomes, we also assessed scenario-specific differences in future projections from 2026 to 2045, assuming continuation of the status quo (i.e. no major changes to current HIV programmes). All estimates are reported as posterior means with 95% credible intervals (CI; 0.025-0.975 quantiles).

### External validation

We evaluated model performance by comparing scenario-specific estimates of HIV incidence, prevalence, and viral load (VL) suppression among FSW with data from the 2019 South African national FSW survey [22,24,29]. This was a large, community-led, cross-sectional survey conducted across all nine provinces. It used an adapted respondent-driven sampling approach to recruit FSW primarily from districts with established sex work programmes. Because incidence and VL suppression were not calibration targets, they served as an independent test of whether the scenarios reproduced observed epidemic patterns beyond the prevalence-based fit.

We also compared scenario-specific modelled age-standardized IRRs between FSW and the general female population with regional estimates for eastern and southern Africa from a recent meta-analysis [8]. Although these pooled IRRs are not specific to South Africa, they provide a useful external benchmark for assessing whether the scenarios capture broader epidemiological patterns seen in similar high-burden settings.

### Sensitivity analysis

As a sensitivity analysis, we further subdivided the “b” scenarios, those in which both FSW age and duration in sex work vary over time. In the additional “b *age only*” and “b *duration only*” scenarios, we allowed only one characteristic to change while holding the other constant.

## Results

All six scenarios achieved good calibration. Most posterior distributions of the 10 or 11 calibrated parameters were similar to prior distributions and across scenarios (Supplementary Table 1). Model fits reproduced the HIV prevalence in the general population and in pregnant women from the calibration data across all scenarios (Supplementary Figure 2 and 3). Estimates for pregnant women and the general population were generally very similar across scenarios. Scenarios with time-varying transmission risk (2a-3b) showed a slightly poorer fit to early ANC data (before 1995) compared with the time-invariant scenarios (1a and 1b).

### Modelled HIV outcomes in sex workers & comparison to validation data

Assuming a constant transmission risk (1a,1b) resulted in the highest modelled HIV incidence and the lowest rate of viral load suppression among sex workers (Figure 2A). In these scenarios, HIV prevalence peaked around 2002 and then declined. By contrast, in the time-varying transmission risk scenarios (2a-3b), prevalence plateaued around 2005 and declined only in the last 5 to 10 years. The two time-varying transmission assumptions led to similar results, although the exposure-dependent changing risk scenarios (3a,3b) generated a steeper rise in HIV incidence early in the epidemic. Allowing FSW age and duration of sex work to increase over time (the “b” scenarios) led to slightly lower HIV incidence and higher HIV prevalence and viral suppression from 2005 onward compared with the time-invariant scenarios (“a”).

**Figure 2.**
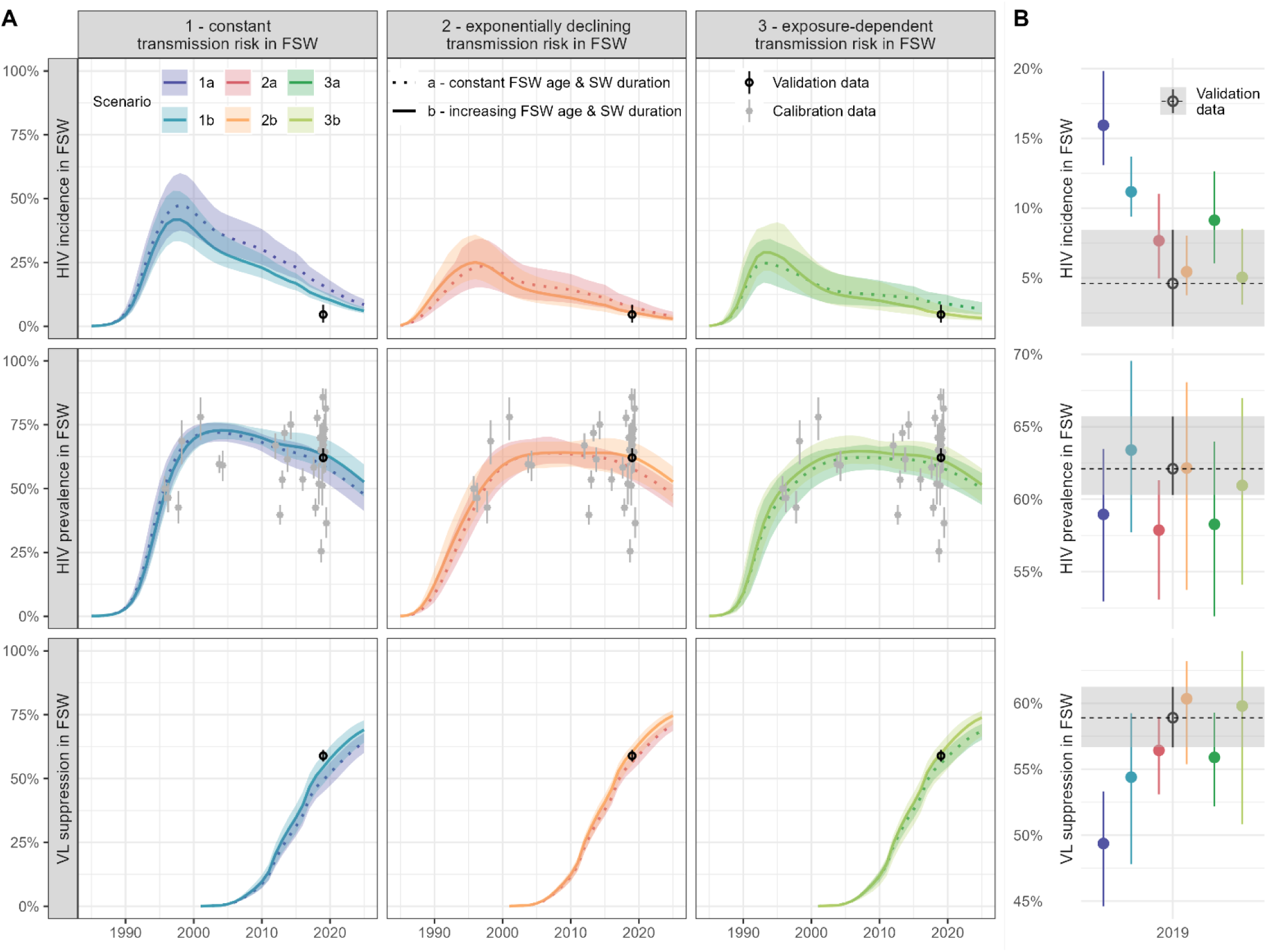
Modelled HIV outcomes among female sex workers (FSW) across six scenarios. Panel A shows modelled time trends in HIV incidence, HIV prevalence, and viral load (VL) suppression among FSW from 1985 to 2025. Columns correspond to different assumptions about HIV transmission risk [(1) constant; (2) exponential decline; (3) exposure-dependent change], and line types distinguish assumptions about FSW characteristics [(a) constant; (b) increasing]. Coloured lines indicate posterior mean estimates, with shaded areas showing 95% credible intervals (2.5th and 97.5th percentiles of the posterior distributions). Gray dots with error bars indicate calibration data for HIV prevalence in FSW. The black dot with error bar indicates the 2019 national FSW survey (validation data [23,25,30]). Panel B zooms into 2019 to facilitate comparison between scenarios and the validation data.

All scenarios reproduced the FSW prevalence calibration data, although the substantial variability in these data made it difficult to determine which scenario fit best. When comparing modelled estimates to the validation data [22,24,29], scenarios 2b and 3b showed the closest agreement, matching the 2019 national survey estimates of HIV incidence (4.6 per 100 pyrs; 95% CI 1.5-8.4), prevalence (62.1%; 60.3-65.7%) and VL suppression (58.9%; 56.7-61.2%) most closely. Modelled estimates for scenario 2b were 5.4 per 100 pyrs (95% CI 3.8-8.0), 62.1% (53.7-68.1%), and 60.3% (55.4-63.2%), while scenario 3b yielded 5.0 per 100 pyrs (CI 3.1-8.5), 60.9% (54.1-67.0%), and 59.8% (50.8-64.0%) (Figure 2B).

### Modelled age-standardized IRR & comparison to meta-regression

Modelled age-standardized IRRs between 1990 and 2025 were consistently above 5 across all scenarios, indicating substantially higher HIV incidence in sex workers than in age-matched women in the general population (Figure 3). Under constant transmission risk (1a,1b), IRRs were initially very high and declined gradually, implying a somewhat faster drop in incidence among sex workers than in the general female population. In the time-varying transmission risk scenarios (2a-3b), IRRs declined steeply from 1990 to 2005, reflecting a much faster early decline in sex workers, but then stabilized. In the exposure-dependent transmission risk scenario with constant FSW characteristics (3a), IRRs rose again after 2005, suggesting slower incidence declines in sex workers relative to the general population. In the post-2005 period, scenarios 2b and 3b were most consistent with the pooled estimates for eastern and southern Africa from the meta-analysis by Jones et al. [8].

**Figure 3.**
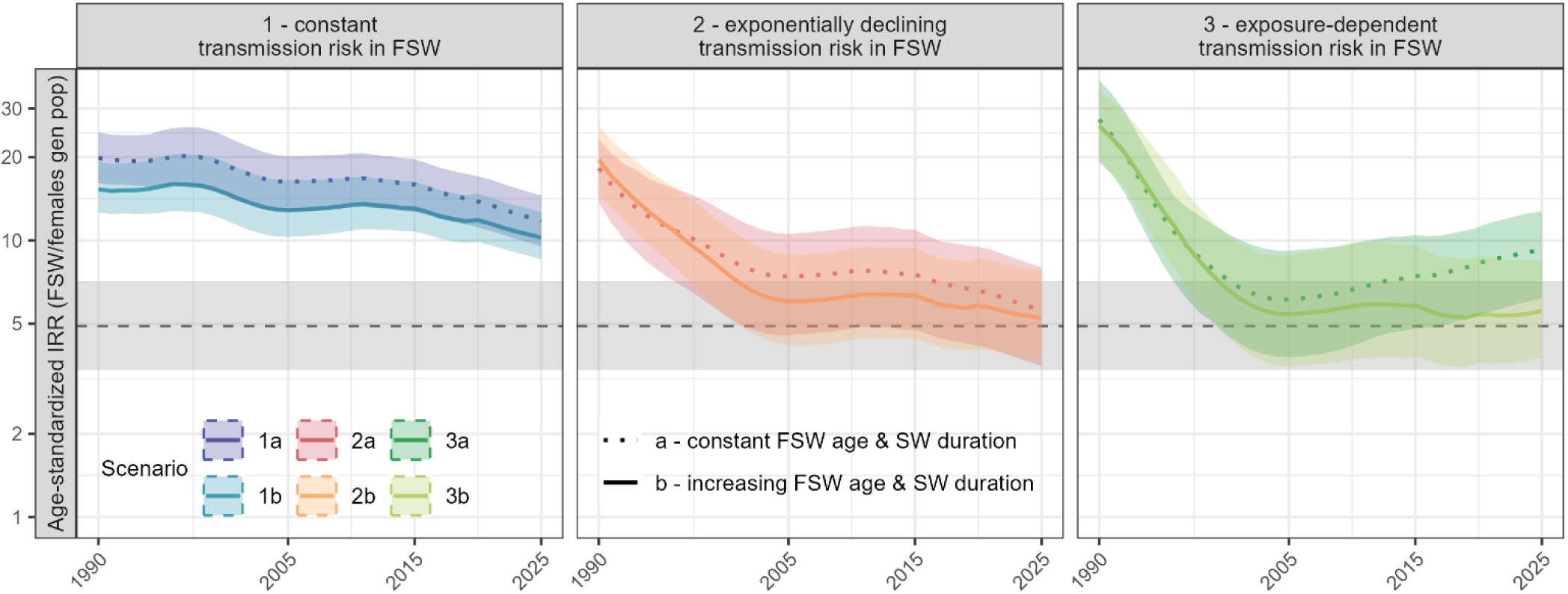
Age-standardized incidence rate ratios (IRRs) comparing HIV incidence in female sex workers (FSW) to women in the general population across six scenarios. Columns correspond to different assumptions about HIV transmission risk [(1) constant; (2) exponential decline; (3) exposure-dependent change], while line types distinguish assumptions about FSW characteristics [(a) constant; (b) increasing]. Coloured lines indicate posterior mean estimates, with shaded areas showing 95% credible intervals (2.5th and 97.5th percentiles of the posterior distributions). The dashed horizontal line (and grey shaded band) indicates the pooled IRR (95% CI) for the eastern and southern Africa region from a recent meta-analysis [2].

### Modelled PAF for commercial sex work

Across scenarios, the estimated PAF of commercial sex work ranged between 7.3% and 22.7% in 2000-2025 (Figure 4). All scenarios showed an initial decline until about 2005, followed by a gradual increase up to 2025, most pronounced under the constant transmission risk assumptions (1a,1b). These scenarios produced the highest PAFs, reaching 22.7% (1a) and 21.0% (1b) in 2025 - almost double the estimates from the time-varying scenarios (12.0% and 10.4% for 2a and 2b; 13.1% and 9.1% for 3a and 3b). Scenarios 2 and 3 resulted in similar estimates up to 2025, but their long-term projections differed: 2a and 2b stabilised with little change, whereas 3b and especially 3a showed a marked increase between 2026 and 2045.

**Figure 4.**
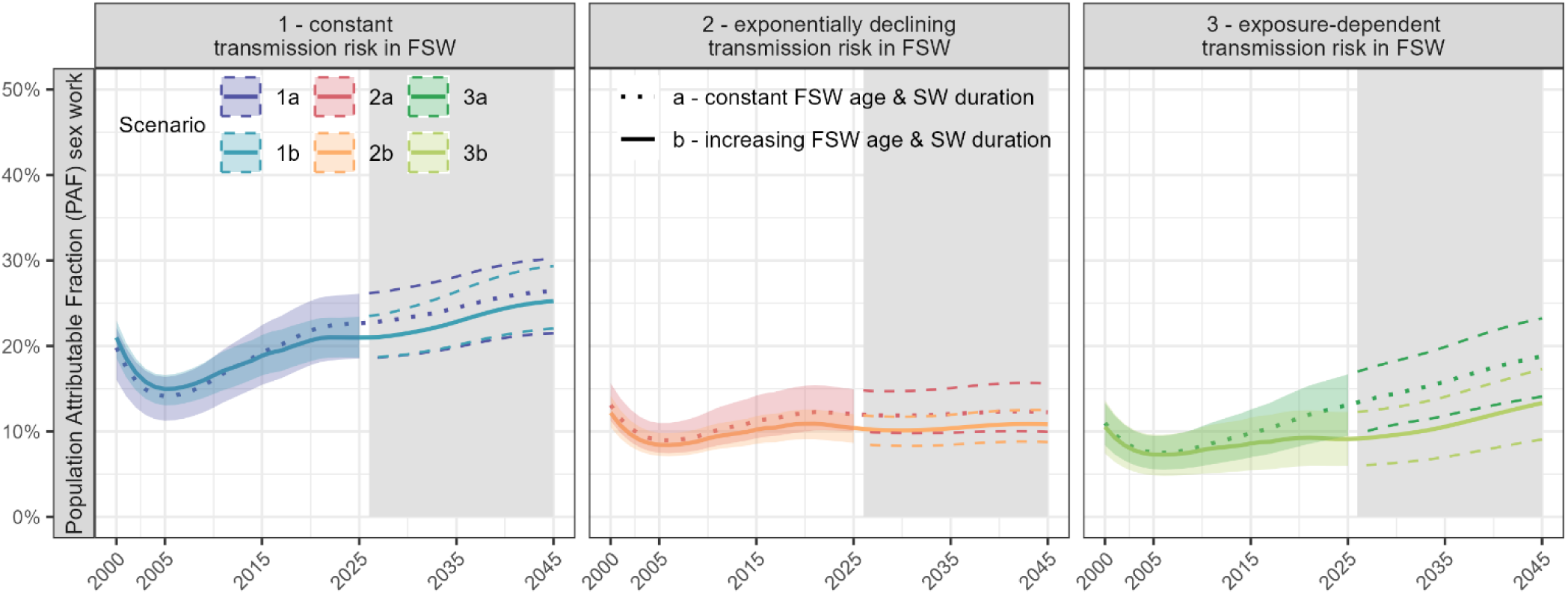
Estimated population attributable fraction (PAF) of commercial sex work, 2000-2045, across scenarios. The PAF is defined as the proportion of yearly new HIV infections in FSW and their clients (restricted to infections acquired through contacts with FSW) among all new adult HIV infections. Columns correspond to different assumptions about HIV transmission risk [(1) constant; (2) exponential decline; (3) exposure-dependent change], while line types distinguish assumptions about FSW characteristics [(a) constant; (b) increasing]. Coloured lines indicate posterior mean estimates, with shaded areas showing 95% credible intervals (2.5th and 97.5th percentiles of the posterior distributions). The grey area marks future projections (2026-2045).

When separating contributions from FSW and their clients, the client-attributable share was higher in all scenarios and increased from 2005 by 1.1 to 1.4-fold, reaching 7-14% in 2025. The share attributable to infections in FSW was lower but increased more steeply, by 1.6 to 3.6-fold to 2-9% in 2025 (Supplementary Figure 4A). The faster relative increase was also reflected in the distribution of sex-work-related infections, where the proportion occurring in FSW increased over time across all scenarios, reaching 18-38% in 2025 (Supplementary Figure 4B). In scenarios 2b and 3b, however, this increase only began around 2020, which marks the start of the period when FSW age and duration of sex work were held constant at 2019 levels in these scenarios.

### Sensitivity analyses

When allowing only one of either FSW age or sex-work duration to increase in the “b” scenarios, modelled HIV estimates showed that their relative impact differed not only by transmission assumption but also by outcome (Supplementary Figure 5). More often, increasing age alone produced results closer to the full “b” assumption than increasing duration alone. For example, under constant transmission risk, HIV incidence estimates with increasing duration only (1b *duration only*) were similar to those with no changes (1a), whereas increasing age only (1b *age only*) yielded results close to the “both increasing” scenario (1b). When compared with the validation data, scenarios 2b and 3b still provided the best agreement with estimates from the 2019 national survey.

## Discussion

### Main findings

Our findings demonstrate that HIV model estimates for sex workers are highly sensitive to assumptions about their characteristics and HIV transmission risk. Models typically treat FSW age, duration of sex work, and per-act client-to-FSW transmission probabilities as constant over time, yet there is evidence that all three parameters have changed substantially. In South Africa, increases in the average age of sex workers and the duration of sex work over the past two decades have been documented [14,15], while data from Kenya provide evidence that HIV transmission probabilities from clients to sex workers were substantially higher early in the epidemic and declined over time [16]. When we incorporated time-varying FSW characteristics and declining transmission risks, the resulting estimates aligned more closely with empirical data on HIV incidence, prevalence, and viral suppression, particularly in scenarios 2b and 3b, which allow both FSW characteristics and transmission risk to vary over time. These scenarios also produced IRRs more consistent with regional estimates from eastern and southern Africa and yielded markedly lower estimates of the contribution of sex work to overall HIV transmission. These results indicate that assuming constant FSW characteristics and transmission risks can overestimate the role of sex work in sustaining the epidemic and may lead to misinformed programme planning and resource allocation.

### Strengths and weaknesses

To the best of our knowledge, this is the first systematic assessment of how common modelling assumptions about sex workers affect HIV estimates. It builds on Thembisa, a national HIV model for South Africa whose structure has been iteratively refined and updated across multiple versions over time. The analysis draws on multiple empirical data sources, including ANC surveillance, national household surveys, and over two decades of FSW survey data, providing a strong empirical basis for model calibration. Independent external validation against the 2019 national FSW survey [22,24,29], together with comparisons to the pooled IRR estimate from other sub-Saharan African countries [8] further strengthens confidence in the findings.

Our study also has some weaknesses. First, assumed trends in FSW age and duration of sex work were informed by our previous synthesis of multiple studies [14], which may not be fully representative of the broader FSW population. Similarly, in the absence of South Africa-specific estimates, assumptions about the early-epidemic decline in client-to-FSW HIV transmission risk were informed by data from a long-standing sex worker cohort in Kenya [16], which may not fully reflect the South African context.

Second, external validation relied on data from the 2019 national FSW survey [22,24,29], which provides the best available national estimate of HIV incidence and prevalence in South Africa. However, because recruitment focused on established sex work programmes and used peer-recruitment methods, the survey may overrepresent sex workers who are more connected to services and therefore have better HIV outcomes than more mobile or hidden sex workers, particularly with respect to viral suppression. Absolute levels of viral suppression estimated from the survey may be biased upwards.

Third, modelled estimates from the early epidemic period (before 2000) should be interpreted with caution, as calibration data for this period are sparse and uncertainty is therefore greater. Projections of future HIV incidence trends also assume a “status quo” scenario, and do not take into account the effect of recent US funding cuts (which have particularly affected FSW programmes [7]) or the potential impact of future interventions, such as the rollout of injectable PrEP [30].

### Interpretation, implications, and comparison with other studies

This study confirms our hypothesis that assuming constant FSW characteristics may lead to underestimating the decline in HIV incidence in sex workers [14]. Allowing for increases in average age and duration of sex work led to a more pronounced decline in HIV incidence after 1995, particularly in the scenarios with time-varying transmission risk. Among the four time-varying transmission risk scenarios, the two that also allowed for increases in FSW age and duration of sex work, and that provided the best fit to the validation data, produced similar results for past and current trends but differed in their future projections, as reflected in differences in predicted PAFs. In the scenario where HIV transmission risk declines early in the epidemic and subsequently remains relatively stable (scenario 2b), the future proportion of HIV transmission associated with sex work (the PAF) is estimated to remain stable. By contrast, when client-to-FSW transmission risk is allowed to vary flexibly with the duration of HIV exposure (scenario 3b), the contribution of sex work to total transmission is projected to increase over the next two decades. This has important implications for FSW-focused programmes, which may justifiably expect a greater share of future HIV funding if the epidemic does indeed become more concentrated in key populations [5,6]. However, our understanding of drivers of heterogeneity in HIV transmission risks among sex workers is still limited [20,21], and further research is needed to support or challenge the assumptions underlying these different models.

Differences in projected PAFs have important implications for how the contribution of sex work to the HIV epidemic is quantified and interpreted. UNAIDS estimated the PAF of sex work for East and Southern Africa to be 16.2% in 2022 [9], based on several models, most of which assume time-invariant FSW parameters [31–34]. In our analysis, the scenario that similarly assumed time-invariant FSW characteristics and transmission risk produced substantially higher PAFs than the best-fitting scenarios 2b and 3b. For example, in 2022, the PAF was estimated at 22.4% under time-invariant assumptions, compared to 10.8% and 9.2% in scenarios 2b and 3b, respectively. Our findings cannot be directly generalized to other models and countries, so this does not mean the UNAIDS estimate should necessarily be revised. Rather, they illustrate how sensitive PAF estimates are to underlying modelling assumptions. Used uncritically, such estimates could understate the effectiveness of existing programmes in reducing HIV incidence and achieving viral suppression in sex workers, leading to an overly pessimistic evaluation of past programme performance.

Of note, the PAFs discussed above capture only direct infections among sex workers and their clients. The transmission population attributable fraction (tPAF) extends this concept by including secondary and onward infections that originate from sex work-related transmission chains. PAF estimates are likely to understate the long-term contribution of sex work to the HIV epidemic [4].

In all scenarios, our model estimates of the temporal IRR trend differed from that in the validation data: the meta-analysis suggested no trend [8], while our model indicated, on average, a long-term decline in IRRs. However, the validation data are based on pooled estimates from multiple countries in eastern and southern Africa and are derived from a meta-regression model, which may lack the statistical power needed to detect a ‘true’ temporal trend. In analyses of cohort-specific data, it was found that the IRR in Zimbabwe increased over the 2009-2019 period [8], roughly consistent with what we found in scenario 3a. It is also worth noting that our model estimates of an overall downward trend in IRRs are driven mainly by reductions in the very early stages of the epidemic (when there was limited incidence data), and our modelled IRRs appear more stable in the period after 2005.

## Conclusions

HIV model estimates for sex workers are highly sensitive to assumptions about trends in FSW characteristics and transmission risk. Relying on constant assumptions may overestimate the contribution of sex work to sustaining the epidemic. Incorporating more realistic, time-varying parameters improves alignment with empirical data. Future HIV modelling efforts should critically assess their assumptions to better capture epidemic dynamics and provide stronger policy guidance for key populations.

## Data Availability

All data produced are available online at

https://github.com/naninatamar/HIV_modelling_in_FSW

## Funding

N.A. was supported by the Swiss National Science foundation (grants P500PM_203010, P5R5PM_225275, PZ00-3_232849). L.J. was supported by the Bill and Melinda Gates Foundation (grant 063625). The conclusions and opinions expressed in this work are those of the authors alone and shall not be attributed to the funders.

## Authors’ contributions

N.A.: Data curation, Formal analysis, Visualisation, Methodology, Writing - original draft, review & editing; M.E.: Writing - original draft, review & editing; K.B.: Writing - review & editing; Y.S.: Writing - review & editing; M.S.: Writing - review & editing; LF.J.: Conceptualisation, Data curation, Methodology, Supervision, Writing - original draft, review & editing

## Availability of data and materials

Scenario-specific implementations of the Thembisa v4.7 C++ model used in this analysis, together with the calibration datasets, model outputs, and R scripts used for data processing and figure generation, are publicly available at: https://github.com/naninatamar/HIV_modelling_in_FSW.

## Ethics approval and consent to participate

This study includes no individual-level data and uses only published information in the public domain. Therefore, ethical approval was not requested.

## Declaration of competing interest

All authors declare no conflict of interest in this study.

## Notes

### Competing Interest Statement

The authors have declared no competing interest.

