## Supplementary material for "Do standard model assumptions realistically represent HIV dynamics in sex workers? A modelling analysis of South African data"

### Supplementary Text: Assumptions implemented in Thembisa

#### (I) Variations in assumptions about sex worker age and duration of sex work.

For the age of sex workers $a(t)$ at time t, Thembisa assumes a Gamma distribution with an offset of 10 years (to prevent unrealistically low ages), and for the duration of sex work $d(t)$, an Exponential distribution. We implemented the following two specifications into Thembisa.

**(a) Time-invariant FSW age & sex work duration.**

$$a(t) - 10 \sim \text{Gamma}(\alpha, \beta), \qquad E[a(t)] = \frac{\alpha}{\beta} + 10 = 29 \text{ and } \text{Var}[a(t)] = \frac{\alpha}{\beta^2} = 9$$

$$d(t) \sim \text{Exp}(\lambda), \qquad E[d(t)] = \frac{1}{\lambda} = 3$$

The values 29 (mean age) and 9 (variance) and 3 (mean duration) come from Thembisa version 4.7 and more details can be found in [1].

**(b) Time-varying FSW age & sex work duration.**

$$a(t) - 10 \sim \text{Gamma}\big(\alpha(t), \beta(t)\big), \qquad E[a(t)] = \frac{\alpha(t)}{\beta(t)} + 10 = \begin{cases} 26.4, & t \leq 1996 \\ \exp\big(3.020 + 0.0133 * (t - 2012.655)\big) + 10, & t \in [1996{,}2019] \\ 32.3, & t \geq 2019 \end{cases} \text{ and CV} = \frac{1}{\sqrt{\alpha(t)}} = 0.34$$

$$d(t) \sim \text{Exp}\big(\lambda(t)\big), \qquad E[d(t)] = \frac{1}{\lambda(t)} = \begin{cases} 2.7, & t \leq 1996 \\ \exp(1.782 + 0.044 \cdot (t - 2013.981)), & t \in [1996, 2019] \\ 7.4, & t \geq 2019 \end{cases}$$

The values 3.020, 0.0133, 2012.655, 0.34 (for FSW age) and 1.782, 0.044, 2013.981 (for duration) are derived from previous work and more details can be found there [2]. The values 26.4, 32.3 (age) and 2.7, 7.4 (duration) correspond to the boundary values of the exponential functions at 1996 and 2019, respectively.

#### (II) Variations in assumptions about HIV transmission risk.

The client-to-FSW transmission probability $\beta_{\text{client}\rightarrow\text{FSW}}(t)$ at time t, Thembisa defines it as a function of the asymptotic (ultimate) transmission probability $\tau$ and additional modifiers. We implemented the following specifications into Thembisa:

**(1) Time-invariant client-to-FSW transmission risk.**

$$\beta_{\text{client}\rightarrow\text{FSW}}(t) = \tau$$

The parameter $\tau$ was estimated during model calibration, assuming a Beta prior (see Supplementary Table 1 for distributional parameters).

**(2) Time-varying client-to-FSW transmission risk (exponential decline).**

$$\beta_{\text{client}\rightarrow\text{FSW}}(t) = \tau \cdot [1 + \big(R_{\text{FSW-start-epidmic}} - 1\big) \cdot 0.75^{t-1985}]$$

Both $\tau$ and $R_{FSW-start-epidmic}$ were estimated during calibration, with Beta and Gamma priors, respectively (see Supplementary Table 1). The factor 0.75 was chosen to produce a rapid transition toward the ultimate rate $\tau$, consistent with the Kenyan study of Kimani et al [3]. See [1] for more details.

**(3) Time-varying client-to-FSW transmission risk (exposure-dependent change).**

$$\beta_{client \to FSW}(t) = \tau \cdot [1 + \left(R_{FSW-start-epidmic} - 1\right) \cdot \exp\left(-4.0 \cdot HIVprevalence_{Clients}(t) \cdot (1 - ARTcoverage_{Clients}(t)) \cdot d(t)\right)]$$

Both $\tau$ and $R_{FSW-start-epidmic}$ were estimated in model calibration and a Beta-prior and Gamma-prior were assumed (see Supplementary Table 1). The factor 4 is chosen such that the decline at the beginning is consistent with the Kenyan study of Kimani et al [3]. HIV prevalence and ART coverage among clients, $HIVprevalence_{Clients}(t)$ and $ARTcoverage_{Clients}(t)$, are computed within each simulation, while the duration $d(t)$ is defined according to the respective duration of sex work assumption described above.

### Supplementary Tables

**Supplementary Table 1. Comparison of prior and posterior distribution of calibrated model parameters.**

Most posterior distributions of the calibrated parameters were similar to their prior distributions and consistent across scenarios, with two notable exceptions. First, for the client-to-FSW transmission probability (τ), the posterior means in scenarios 1a and 1b lay outside the 95% credible interval of the prior distribution. These higher transmission probabilities likely reflect the model's difficulty in reproducing the rapid increase in FSW HIV prevalence during the early epidemic without either (i) an explicit early-epidemic multiplier on the transmission probability or (ii) a higher overall mean transmission probability. Second, for the annual rate of FSW contacts, posterior means in scenarios 1a and 1b were close to or outside the 95% prior credible interval, suggesting that matching the rapid epidemic growth observed in the 1990s requires relatively high FSW–client transmission rates. More information on the different parameters are provided in [1].

| Parameter | Prior distribution | Prior mean (95% CI) | Posterior mean (95% CI) Scenario 1a | Scenario 1b | Scenario 2a | Scenario 2b | Scenario 3a | Scenario 3b |
|---|---|---|---|---|---|---|---|---|
| Gamma density of relative rates of short-term partnership formation by age, in unmarried females | | | | | | | | |
| Mean | Gamma(49.0, 1.40) | 35.0 (25.9-45.5) | 32.2 (30.4-33.9) | 32.2 (30.7-34.1) | 35.1 (33.1-37.0) | 34.6 (33.1-36.0) | 35.4 (33.4-37.4) | 35.7 (33.7-38.1) |
| Standard deviation | Gamma(121.0, 5.5) | 22.0 (18.3-26.1) | 20.7 (19.2-22.3) | 22.1 (20.2-24.3) | 21.8 (19.5-24.5) | 22.7 (21.2-24.3) | 21.8 (19.4-25.2) | 22.7 (20.5-25.4) |
| Sexual mixing parameter | Beta(5.80, 3.87) | 0.600 (0.295-0.866) | 0.737 (0.590-0.840) | 0.763 (0.654-0.858) | 0.623 (0.536-0.700) | 0.792 (0.729-0.858) | 0.608 (0.499-0.677) | 0.660 (0.552-0.766) |
| Female-to-male transmission probability in short-term/ non-spousal partnerships | Beta(7.05, 874) | 0.0080 (0.0032-0.0149) | 0.0064 (0.0059-0.0071) | 0.0063 (0.0055-0.0071) | 0.0068 (0.0061-0.0076) | 0.0071 (0.0066-0.0078) | 0.0068 (0.0061-0.0075) | 0.0070 (0.0064-0.0077) |
| Male-to-female transmission probability in short-term/ non-spousal partnerships | Beta(5.68, 468) | 0.0120 (0.0043-0.0236) | 0.0108 (0.0099-0.0121) | 0.0105 (0.0094-0.0114) | 0.0115 (0.0101-0.0129) | 0.0106 (0.0098-0.0116) | 0.0117 (0.0108-0.0132) | 0.0114 (0.0102-0.0129) |
| RR HIV+ fertility before diagnosis/immune decline | Gamma(100, 76.9) | 1.30 (1.06-1.57) | 1.31 (1.23-1.39) | 1.32 (1.24-1.40) | 1.29 (1.19-1.39) | 1.28 (1.20-1.35) | 1.28 (1.19-1.38) | 1.26 (1.18-1.34) |
| Initial HIV prevalence in high-risk women, ages 15-49 | Uniform(0, 0.001) | 0.050% (0.003-0.098%) | 0.069% (0.044-0.088%) | 0.074% (0.051-0.092%) | 0.063% (0.033-0.087%) | 0.058% (0.025-0.081%) | 0.073% (0.034-0.095%) | 0.072% (0.021-0.096%) |
| Annual rate of FSW contact for unmarried high risk men aged 21 | Gamma(5.444, 1.555) | 3.50 (1.21-6.99) | 6.67 (4.79-8.48) | 7.51 (5.92-8.85) | 4.88 (3.74-6.09) | 4.26 (3.35-5.28) | 4.20 (2.86-5.61) | 3.85 (2.14-5.64) |
| Proportionate reduction in entry into sex work after HIV diagnosis | Uniform(0,1) | 0.500 (0.025-0.975) | 0.582 (0.387-0.766) | 0.725 (0.424-0.896) | 0.349 (0.156-0.561) | 0.383 (0.121-0.683) | 0.361 (0.161-0.624) | 0.536 (0.257-0.864) |
| Client-to-FSW transmission probability per sex act (τ) | Beta(3.995, 3991) | 0.0010 (0.0003-0.0022) | 0.0028 (0.0022-0.0037) | 0.0026 (0.0021-0.0032) | 0.0013 (0.0009-0.0019) | 0.0014 (0.0010-0.0020) | 0.0011 (0.0007-0.0016) | 0.0012 (0.0008-0.0022) |
| Relative susceptibility of FSWs at start of epidemic compared to 1995 ($R_{FSW-start-epidmic}$) | Gamma(6.25, 1.25) | 5.00 (1.88-9.62) | - | - | 5.53 (3.13-7.95) | 7.11 (5.28-9.61) | 4.86 (2.84-7.45) | 5.00 (3.47-7.23) |

### Supplementary Figures

**Supplementary Figure 1. Different assumptions implemented in Thembisa.** Panel A shows the assumed distributions for female sex worker (FSW) characteristics (FSW age and sex work duration). Lines indicate means; shaded areas show the central 50% and 95% intervals. Panel B shows the client-to-FSW transmission risk; lines indicate *posterior* means and shaded areas the 50% and 95% credible intervals. Posterior distributions of "a" vs "b" scenarios differ slightly due to calibration; in Scenarios 3a/3b the parameter additionally depends on the sex work duration (which differs between 3a and 3b – see panel A), hence the larger differences in the posterior distributions.

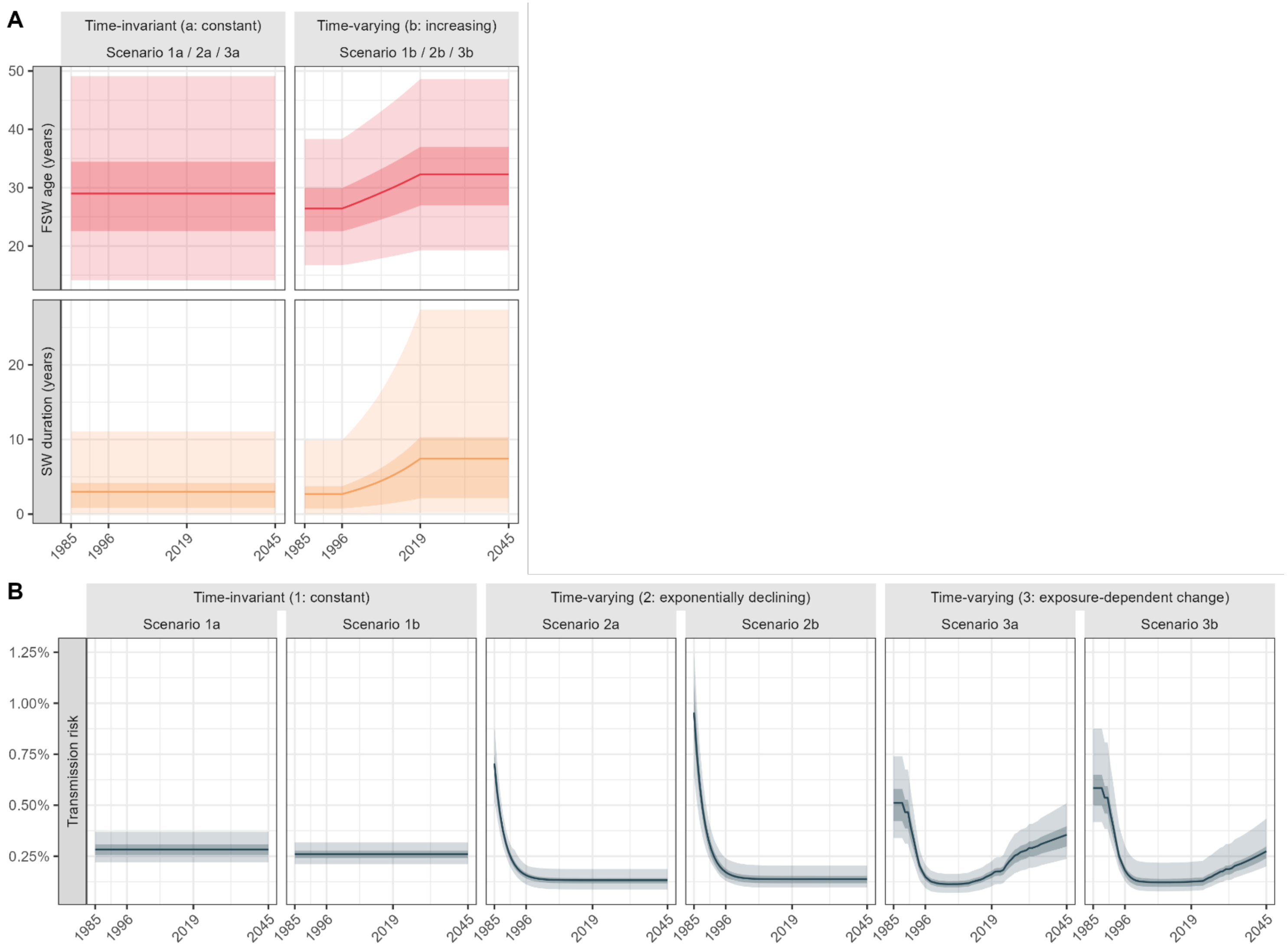

**Supplementary Figure 2. Calibration of the different scenarios to the antenatal survey HIV prevalence data.**

Columns correspond to different assumptions about HIV transmission risk [(1) constant; (2) exponential decline; (3) exposure-dependent change], while line types distinguish assumptions about FSW characteristics [(a) constant; (b) increasing]. Coloured lines indicate posterior mean estimates, with shaded areas showing 95% credible intervals (2.5th and 97.5th percentiles of the posterior distributions). Black dots and error bars represent HIV prevalence levels in pregnant women attending antenatal clinics reported in surveys conducted from 1990-2022.

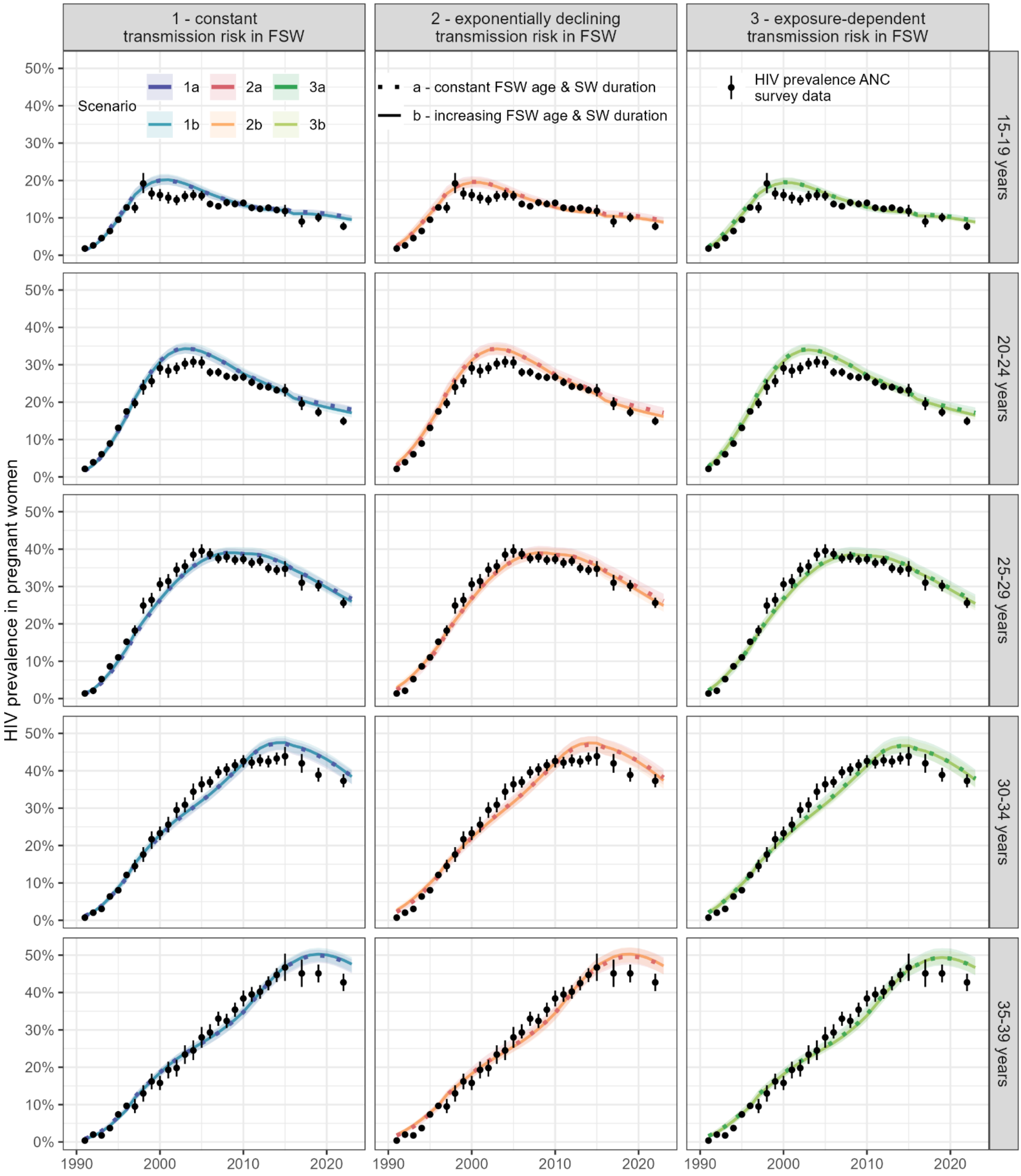

**Supplementary Figure 3. Calibration of the different scenarios to HIV prevalence in the general population.**

Black dots and error bars represent HSRC (Human Sciences Research Council) survey prevalence estimates, together with 95% confidence intervals by age group, stratified by sex (columns) and survey year (rows). Coloured dots and error bars show the posterior mean and 95% confidence intervals of the estimates across scenarios.

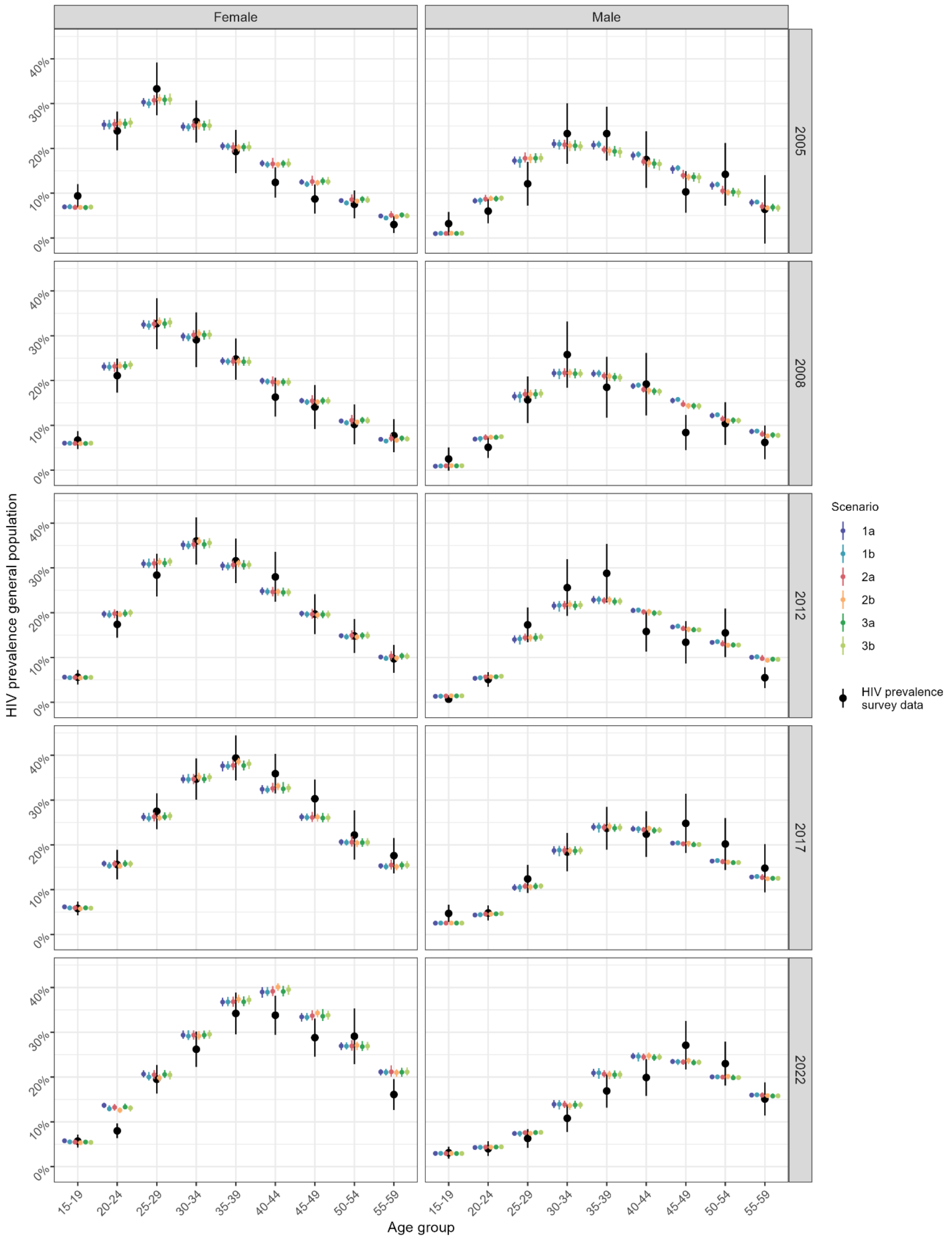

**Supplementary Figure 4. Disaggregated contributions of sex workers and clients, 2000-2045, across scenarios.**

Columns correspond to different assumptions about HIV transmission risk [(1) constant; (2) exponential decline; (3) exposure-dependent change], while line types distinguish assumptions about FSW characteristics [(a) constant; (b) increasing]. Coloured lines indicate posterior mean estimates, with shaded areas showing 95% credible intervals (2.5th and 97.5th percentiles of the posterior distributions). The grey area marks future projections (2026-2045). Panel A shows the PAF of commercial sex work disaggregated by FSW (first row; new HIV infections in FSW / total new adult HIV infections) and clients (second row; new HIV infections in clients due to contact with FSW / total new adult HIV infections). Panel B shows the share of all sex-work-related infections occurring in FSW, calculated as new infections in FSW divided by the sum of new infections in FSW and in clients due to contact with FSW.

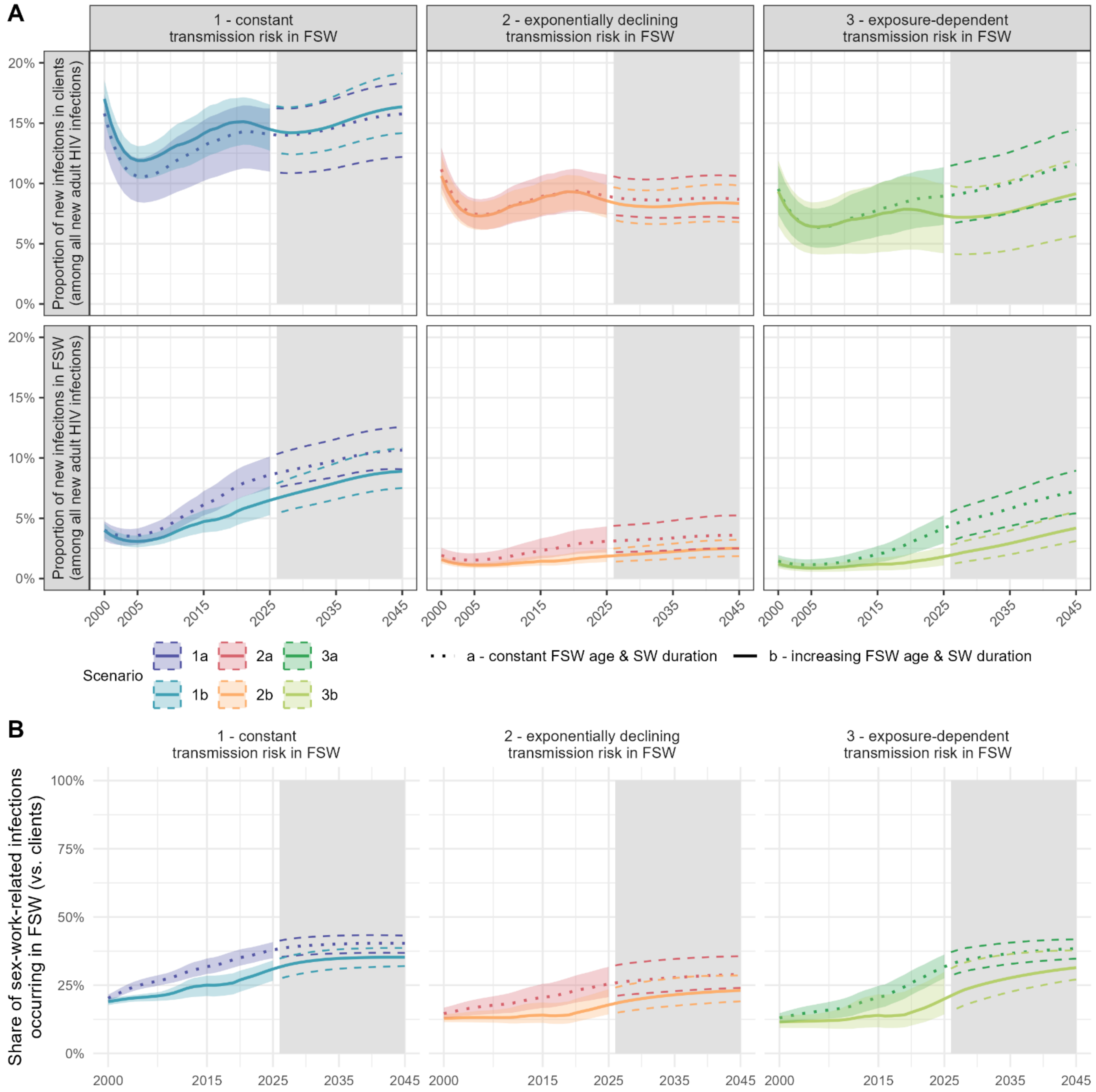

**Supplementary Figure 5. Sensitivity analyses: "b" scenarios separated into changing female sex worker age only versus changing sex work duration only.**
Modelled time trends in HIV incidence, HIV prevalence, and viral load (VL) suppression among FSW from 1985 to 2025. Columns correspond to different assumptions about HIV transmission risk [(1) constant; (2) exponential decline; (3) exposure-dependent change]. Black lines show the modelled estimates under assumptions from the main analysis [(a) constant FSW age & SW duration - dotted line; (b) increasing FSW age & SW duration - solid line]. Dashed coloured lines show modelled estimates for allowing only FSW age (pink) or SW duration (orange) to increase over time in the "b" scenarios. For better visibility only posterior means without 95% credible intervals are shown. Gray dots with error bars indicate calibration data for HIV prevalence in FSW. The black dot with error bar indicates the 2019 national FSW survey (validation data [23,25,30].

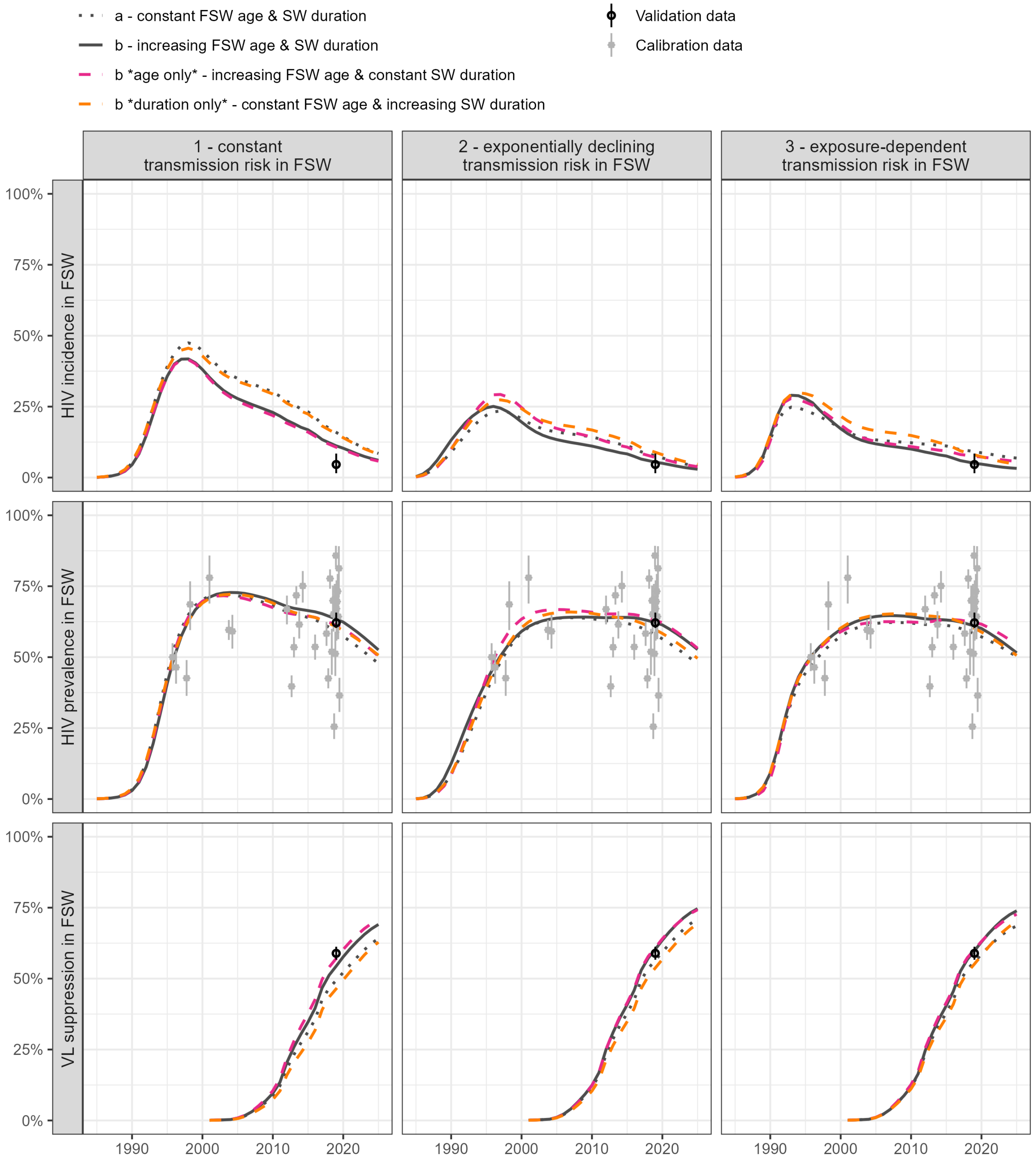
